# Vaccine Effectiveness of Primary Series and Booster Doses against Omicron Variant COVID-19-Associated Hospitalization in the United States

**DOI:** 10.1101/2022.06.09.22276228

**Authors:** Katherine Adams, Jillian P. Rhoads, Diya Surie, Manjusha Gaglani, Adit A. Ginde, Tresa McNeal, Shekhar Ghamande, David Huynh, H. Keipp Talbot, Jonathan D. Casey, Nicholas M. Mohr, Anne Zepeski, Nathan I. Shapiro, Kevin W. Gibbs, D. Clark Files, Madeline Hicks, David N. Hager, Harith Ali, Matthew E. Prekker, Anne E. Frosch, Matthew C. Exline, Michelle N. Gong, Amira Mohamed, Nicholas J. Johnson, Vasisht Srinivasan, Jay S. Steingrub, Ithan D. Peltan, Samuel M. Brown, Emily T. Martin, Arnold S. Monto, Adam S. Lauring, Akram Khan, Catherine L. Hough, Laurence W. Busse, Caitlin C. ten Lohuis, Abhijit Duggal, Jennifer G. Wilson, Alexandra June Gordon, Nida Qadir, Steven Y. Chang, Christopher Mallow, Carolina Rivas, Hilary M. Babcock, Jennie H. Kwon, James D. Chappell, Natasha Halasa, Carlos G. Grijalva, Todd W. Rice, William B. Stubblefield, Adrienne Baughman, Christopher J. Lindsell, Kimberly W. Hart, Sandra N. Lester, Natalie J. Thornburg, SoHee Park, Meredith L. McMorrow, Manish M. Patel, Mark W. Tenforde, Wesley H. Self

**Author notes:** For the Influenza and Other Viruses in the Acutely Ill (IVY) Network***. Katherine Adams, Jillian P. Rhoads, and Diya Surie contributed equally to this work as lead author. Wesley H. Self and Mark W. Tenforde contributed equally to this work as senior author. A full list of collaborators in the IVY Network is shown in Appendix A in the Supplemental Materials. Disclaimer: The findings and conclusions in this report are those of the authors and do not necessarily represent the official position of the Centers for Disease Control and Prevention (CDC). Corresponding Author: Wesley H. Self, MD, MPH; Vanderbilt University Medical Center; Nashville, Tennessee USA. Second Corresponding Author: Katherine Adams, MPH; Influenza Division, Centers for Disease Control and Prevention, 1600 Clifton Rd, MS H24-7, Atlanta, Georgia 30329.

## Abstract

**Objectives:** To compare the effectiveness of a primary COVID-19 vaccine series plus a booster dose with a primary series alone for the prevention of Omicron variant COVID-19 hospitalization.

**Design:** Multicenter observational case-control study using the test-negative design to evaluate vaccine effectiveness (VE).

**Setting:** Twenty-one hospitals in the United States (US).

**Participants:** 3,181 adults hospitalized with an acute respiratory illness between December 26, 2021 and April 30, 2022, a period of SARS-CoV-2 Omicron variant (BA.1, BA.2) predominance. Participants included 1,572 (49%) case-patients with laboratory confirmed COVID-19 and 1,609 (51%) control patients who tested negative for SARS-CoV-2. Median age was 64 years, 48% were female, and 21% were immunocompromised; 798 (25%) were vaccinated with a primary series plus booster, 1,326 (42%) were vaccinated with a primary series alone, and 1,057 (33%) were unvaccinated.

**Main Outcome Measures:** VE against COVID-19 hospitalization was calculated for a primary series plus a booster and a primary series alone by comparing the odds of being vaccinated with each of these regimens versus being unvaccinated among cases versus controls. VE analyses were stratified by immune status (immunocompetent; immunocompromised) because the recommended vaccine schedules are different for these groups. The primary analysis evaluated all COVID-19 vaccine types combined and secondary analyses evaluated specific vaccine products.

**Results:** Among immunocompetent patients, VE against Omicron COVID-19 hospitalization for a primary series plus one booster of any vaccine product dose was 77% (95% CI: 71–82%), and for a primary series alone was 44% (95% CI: 31–54%) (p<0.001). VE was higher for a boosted regimen than a primary series alone for both mRNA vaccines used in the US (BNT162b2: primary series plus booster VE 80% (95% CI: 73-85%), primary series alone VE 46% (95% CI: 30-58%) [p<0.001]; mRNA-1273: primary series plus booster VE 77% (95% CI: 67-83%), primary series alone VE 47% (95% CI: 30-60%) [p<0.001]). Among immunocompromised patients, VE for a primary series of any vaccine product against Omicron COVID-19 hospitalization was 60% (95% CI: 41-73%). Insufficient sample size has accumulated to calculate effectiveness of boosted regimens for immunocompromised patients.

**Conclusions:** Among immunocompetent people, a booster dose of COVID-19 vaccine provided additional benefit beyond a primary vaccine series alone for preventing COVID-19 hospitalization due to the Omicron variant.

## BACKGROUND

The highly transmissible SARS-CoV-2 Omicron variant, first identified in southern Africa in November 2021, became the dominant variant in the United States (US) by late December 2021.^1,2^ The surges in COVID-19 cases driven by this variant and its emergent lineages (PANGO BA.1/BA.2) was the largest recognized in the United States, with a peak of over 400,000 cases per day.^3^ Emerging data on vaccine protection against disease caused by the Omicron variant has been mixed. Early immunological studies showed evidence of immune evasion by the Omicron variant with decreased neutralization from either vaccine sera or monoclonal antibodies compared to previous variants.^4,5^ Recent evidence suggests a rapid decline over time in antibody titers following booster doses.^6^ Real-world studies have shown decreased effectiveness of both primary series and booster doses of COVID-19 vaccine regimens against symptomatic infection, with waning occurring as early as 5–9 weeks after booster dose receipt.^7–9^ However, results from studies evaluating the protection provided by booster doses against severe COVID-19 due to the Omicron variant have varied, with some studies suggesting robust protection similar to protection against earlier SARS-CoV-2 variants, and others suggesting reduced protection against the Omicron variant and further reductions over time as the time from booster dosing increases.^8,10–14^ Additionally, questions remain about the effectiveness of booster vaccine doses and whether specific demographic and high-risk groups benefit most from COVID-19 booster doses.^15^

Understanding the effectiveness of COVID-19 vaccines against severe disease caused by the Omicron variant and its lineages has been challenging, which likely plays a role in the variability of recent results that have emerged. Many hospitalized patients tested positive for SARS-CoV-2 in the first quarter of 2022; however, in part due to the high incidence of infections caused by the Omicron variant and universal testing upon admission, some fraction of these detections were likely “incidental” in patients admitted for alternative reasons and may not have represented hospitalizations due to COVID-19.^16,17^ If VE is lower against milder infections compared with severe infections, incidental SARS-CoV-2 detections may artificially reduce VE estimates against severe COVID-19, when using any COVID-19 detections among hospitalized patients as an outcome.^10^ Increased time since vaccination with a primary series and widespread use of booster vaccine doses during the Omicron period further complicate distinguishing the contribution of immune evasion of the Omicron variant from waning immunity.^8,10^

To overcome these challenges, we evaluated the effectiveness of COVID-19 vaccines to prevent hospitalizations during the Omicron predominant period in the US, using data from patients hospitalized primarily because of COVID-19. We calculated vaccine effectiveness (VE) of a primary series plus one booster dose and a primary vaccine series alone, by time since the last vaccine dose, thus providing data to help understand the effectiveness of booster doses and the interplay of immune evasion and waning immunity during the Omicron surge. Lastly, as a complementary laboratory analysis to these clinical VE evaluations, we measured serum antibody concentrations in healthy adult volunteers to determine anti-SARS-CoV-2 responses before and after receiving booster doses.

## METHODS

### Setting and Design for VE Analysis

This prospective, multicenter observational assessment was conducted by the Influenza and Other Viruses in the Acutely Ill (IVY) Network in collaboration with the US Centers for Disease Control and Prevention (CDC). The IVY Network is a 21-hospital collaborative in the US that has published COVID-19 VE estimates iteratively throughout the pandemic (**Table S1**). The current analysis included adult patients hospitalized at IVY sites during the period of Omicron variant predominance in the Network: December 26, 2021–April 30, 2022 (with a brief pause of enrollment January 25–31, 2022 for a protocol update). Using a test-negative design, VE for the prevention of Omicron-variant COVID-19-associated hospitalization was calculated for the COVID-19 vaccines authorized for use in US: BNT162b2 (Pfizer-BioNTech), mRNA-1273 (Moderna), and Ad26.COV2 (Johnson & Johnson [Janssen]).^18–20^ These activities were reviewed by CDC, were conducted consistent with applicable federal law and CDC policy (45 C.F.R. part 46.102(l)(2), 21 C.F.R. part 56; 42 U.S.C. §241(d); 5 U.S.C. §552a; 44 U.S.C. §3501 et seq), and were determined to be public health surveillance with waiver of informed consent by institutional review boards at CDC and each enrolling site.

### Participants

Site personnel prospectively identified and enrolled hospitalized COVID-19 case-patients and concurrent test-negative control-patients. Case-patients were adults (≥18 years old) admitted to the hospital with symptomatic COVID-19 confirmed with a positive SARS-CoV-2 reverse transcription-polymerase chain reaction (RT-PCR) or antigen test within 14 days of symptom onset. Case-patients had one or more of the following COVID-19-associated signs or symptoms: fever, cough, shortness of breath, loss of taste, loss of smell, use of respiratory support (high flow oxygen by nasal cannula, non-invasive ventilation, or invasive mechanical ventilation) for the acute illness, or new pulmonary findings on chest imaging indicating pneumonia. Control-patients were adults hospitalized with at least one of the above signs or symptoms of COVID-like illness who tested negative for SARS-CoV-2, influenza, and respiratory syncytial virus by RT-PCR within 14 days of symptom onset. Case-control status was determined by results of both clinical SARS-CoV-2 testing at the admitting hospital as well as standardized, central laboratory RT-PCR SARS-CoV-2 testing. Patients who tested positive for SARS-CoV-2 by either clinical testing or central laboratory testing were classified as cases, while patients who tested negative by both clinical and central laboratory testing were classified as controls. Sites sought to enroll case and control-patients in a 1:1 ratio. Case- and control-patients were enrolled within two weeks of one another and not matched on other individual patient characteristics.

### Data Collection

Trained personnel at enrolling sites collected patient data on demographics, medical history, underlying health conditions (**Table S2**), COVID-19 vaccination status, laboratory findings, and clinical outcomes through patient or proxy interviews and medical record review. Patients were followed from admission until discharge, death, or hospital day 28 (whichever occurred first). Clinical outcomes included in-hospital death, invasive mechanical ventilation, supplemental oxygen therapy, vasopressor use, and intensive care unit admission. The composite outcome of death or invasive mechanical ventilation was used to identify patients with “critical COVID-19.” Individual pre-existing health conditions were grouped into condition categories of cardiovascular, endocrine, gastrointestinal, hematologic, immunocompromising, neurologic, pulmonary, and renal. Verification of COVID-19 vaccination was performed for information such as dates of vaccination, vaccine products, and lot numbers using a systematic search of hospital electronic medical records, state vaccine registries, and vaccination cards (when available).

### Classification of Vaccination Status

We assessed effectiveness of a primary COVID-19 vaccine series plus one booster dose and a primary series alone. During the surveillance period, US recommendations for COVID-19 vaccine dosing for primary series and booster doses varied depending on health status (notably, presence of immunocompromising conditions), age, vaccine product, and timing of prior vaccination. While national US vaccine surveillance data do not distinguish vaccination status by medical conditions, the IVY network collects detailed clinical data to facilitate descriptions of vaccine uptake by nuanced eligibility criteria.^3^ Based on CDC recommendations, we classified patients into four mutually exclusive vaccination status groups: 1) unvaccinated; 2) partially vaccinated; 3) primary series alone; 4) primary series plus one booster dose (**Table S3**).

Among immunocompetent individuals (i.e., those without an immunocompromising condition collected by the IVY network [**Table S2**]), we classified patients as completing a primary series if they received either one dose of Ad26.COV2 (Janssen) or two doses (>3-8 weeks apart) of an mRNA vaccine (BNT162b2 or mRNA-1273) with the final dose ≥14 days before illness onset.^22^ Immunocompetent patients were classified as having received a booster dose if they completed a primary series and received an additional dose of any licensed COVID-19 vaccine ≥7 days prior to illness onset.

Among immunocompromised individuals, we classified patients as completing a primary series if they received either an initial Ad26.COV2 dose plus one additional COVID-19 vaccine dose (AD26.COV2 or mRNA) or three doses of an mRNA vaccine ≥7 days before illness onset. Immunocompromised patients were classified as receiving a booster dose if they completed a primary series and received an additional dose of any licensed COVID-19 vaccine product ≥7 days prior to illness onset.

Patients were classified as unvaccinated if they had never received any COVID-19 vaccine. Those who received one mRNA COVID-19 vaccine dose but did not complete a primary series were classified as partially vaccinated and excluded from this analysis. Sample size of patients who receive two booster doses was not large enough to calculate VE for a primary series plus two booster doses; thus, patients who received two booster doses were excluded from this analysis. Patients were also excluded if they received a booster dose prior to eligibility based on US recommendations (**Table S4**).

### Molecular diagnosis and sequencing

Upper respiratory specimens were obtained from enrolled patients by nasal swab or saliva collection for SARS-CoV-2 testing. The samples underwent standardized RT-PCR testing at a central laboratory at Vanderbilt University Medical Center (Nashville, Tennessee) for detection of two SARS-CoV-2 nucleocapsid gene targets (N1 and N2). Samples with detection of either N1 or N2 with a cycle threshold ≤32 were shipped to the University of Michigan (Ann Arbor, Michigan) for viral whole genome sequencing using the ARTIC Network protocol (v4.1 primer set) and an Oxford Nanopore Technologies GridION instrument.^23^ SARS CoV-2 variants and lineages are reported using the PANGO nomenclature.^24^ Case-patients with a SARS-CoV-2 variant other than Omicron identified through sequencing were excluded from this analysis.

### Statistical Analysis

VE for the prevention of COVID-19-associated hospitalization was estimated by comparing the odds of antecedent COVID-19 vaccination versus no vaccination among case- and control-patients.^20^ Adjusted odds ratios (aORs) were generated using multivariable logistic regression models with case status as the outcome, vaccination as the primary exposure, and prespecified covariables considered potential confounders: admission date (biweekly intervals), age (18–49, 50–64, and ≥65 years), sex, self-reported race and ethnicity, and U.S. Health and Human Services region of the admitting hospital. VE was calculated as (1 – aOR) x 100.

Models were stratified by vaccination group (primary series plus one booster dose or primary series alone), immunosuppression status (immunocompetent or immunocompromised), age group (18– 64 versus ≥65 years), time from receipt of last vaccine dose to illness onset, and vaccine product received (Ad26.COV2, BNT162b2, or mRNA-1273). Time between last vaccine dose of a primary series and illness onset was stratified into 14–150 days and >150 days to align with current US recommendations for the timing of a first booster dose.^25^ Time between first booster dose and illness onset was dichotomized into 7–120 days and >120 days based on timing of eligibility for a second booster dose as recently allowed in select populations.^26^

Statistical differences among baseline characteristics, outcomes, and treatments were evaluated using Pearson’s chi-squared test for binary or categorical variables and the Wilcoxon rank-sum test for continuous variables. Vaccine effectiveness P values were calculated by fitting a multilevel regression model and performing a pairwise comparison across levels of categorical variables (vaccination group, vaccine product). Statistical analyses were performed using Stata 16 (College Station, TX) and GraphPad Prism version 9.3.1 for Windows (GraphPad Software, San Diego, CA).

### Post-vaccination antibody responses

To complement the VE analysis described above, we also evaluated anti-SARS-CoV-2 antibody responses after booster doses with the BNT162b2, mRNA-1273, and Ad26.COV2 vaccine products. Healthy adult volunteers scheduled for their first booster dose were recruited at four IVY Network sites during October 5, 2021–January 28, 2022. Enrolled participants reported never having symptomatic COVID-19, either at the time of enrollment or in the past, and never testing positive for SARS-CoV-2.

Serum was collected before the booster dose and then again 2–6 weeks after the dose. These serum samples were tested at CDC with the V-PLEX SARS-CoV-2 panel 2 kit (Meso Scale Diagnostics) for IgG against the spike protein (S), receptor binding domain (RBD), and nucleocapsid (N). Targets of the antibody responses were based on proteins from the USA-WA1/2020 strain. Participants with elevated anti-nucleocapsid antibody concentration (>11.8 binding antibody units [BAU] per mL) suggestive of a prior SARS-CoV-2 infection at either the pre-booster or post-booster time point were excluded. Pre-booster and post-booster anti-S and anti-RBD concentrations were summarized with spaghetti plots and geometric means. Some participants had previously contributed serum specimens for antibody measurements 2–6 weeks after a primary series (i.e., after a second mRNA dose or after a first Ad26.COV2 dose) earlier in this program.^27^ When available, these post-primary series antibody measurements were displayed to facilitate evaluation of antibodies in the same individual after a primary series, before a first booster dose, and after a first booster dose.

### Patient and public involvement

Patients were not involved in the original design of this study. Throughout the conduct of the study, patients were routinely engaged in the work via structured interviews conducted between research personnel and patients, which included discussions about COVID-19 and COVID-19 vaccines. Results of the study will be disseminated to the public via announcements from the Centers for Disease Control and Prevention and via lay and social media.

## RESULTS

### Participants in VE Analyses

During December 26, 2021–April 30, 2022, 4,299 hospitalized patients were enrolled from 21 hospitals. 1,118 (26%) patients were excluded from the analysis, with the most common reason being partial vaccination (n=461) (**Figure S1**). 3,181 patients were included in the final analysis, including 1,572 (49%) COVID-19 case-patients and 1,609 (51%) test-negative control-patients, with a median overall age of 64 years (IQR 52–74). 1,520 (48%) included patients were female, 619 (19%) were non-Hispanic Black, 423 (13%) were Hispanic, and 654 (21%) were immunocompromised (**Table 1)**.

**Table 1.**
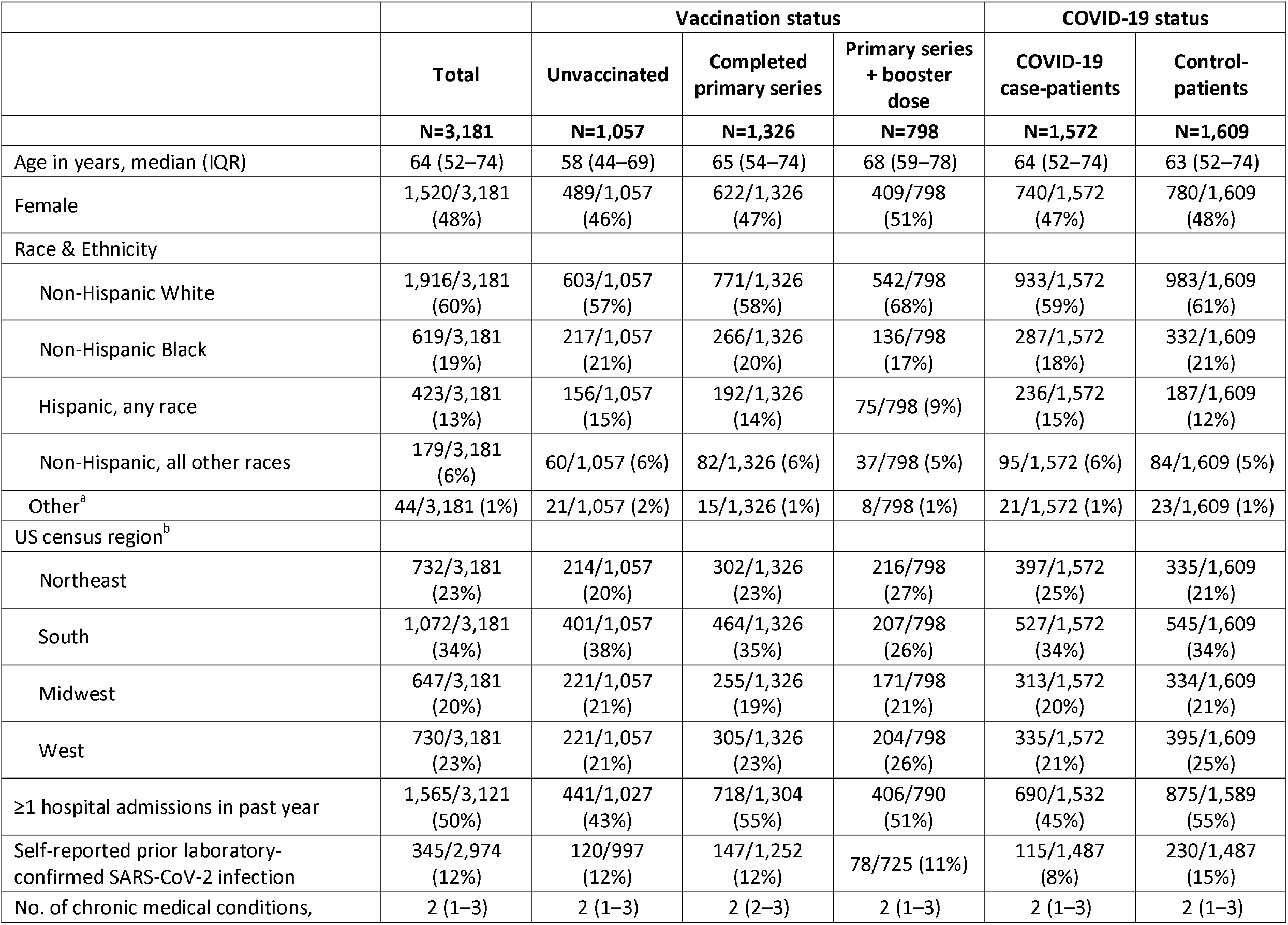
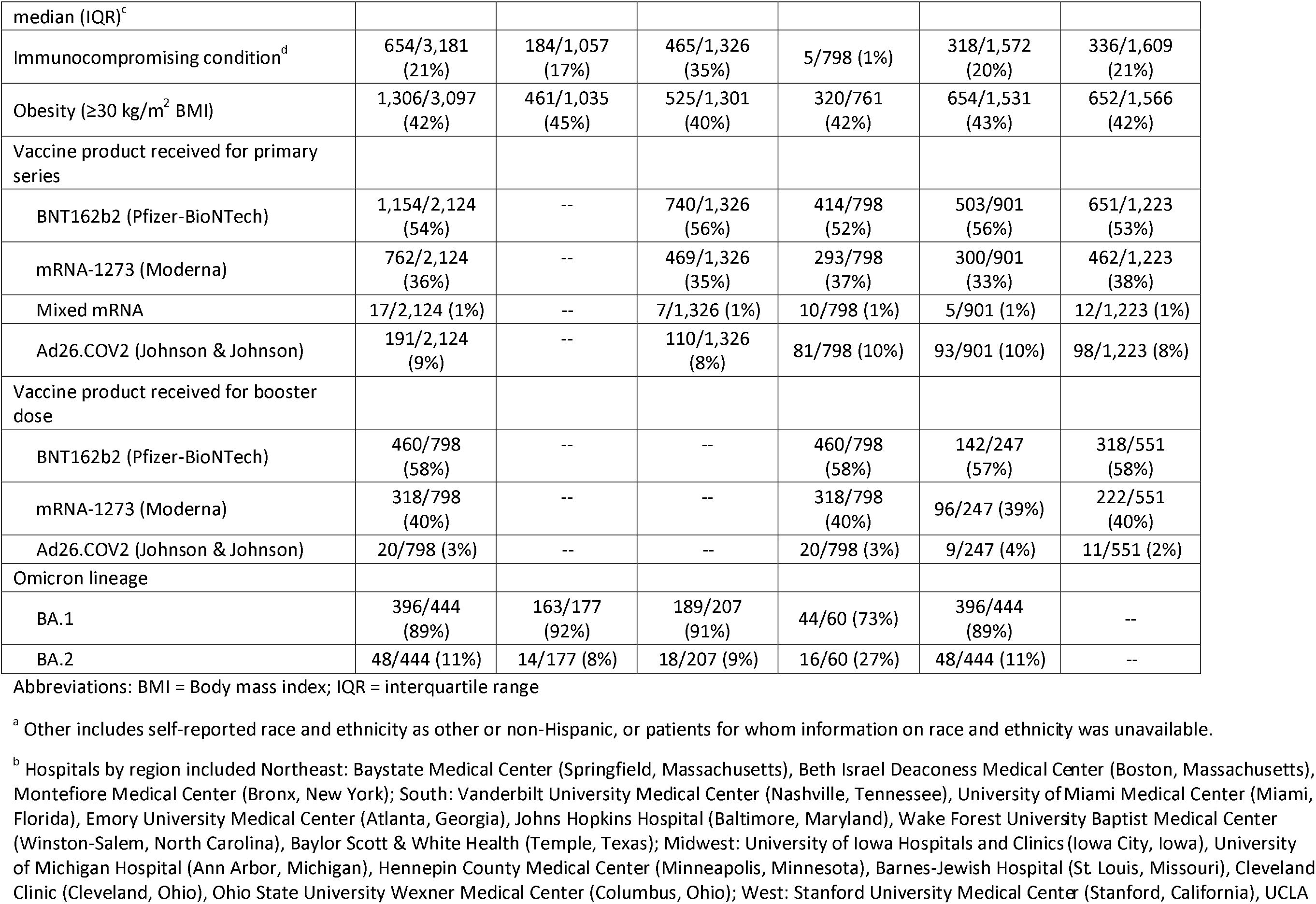
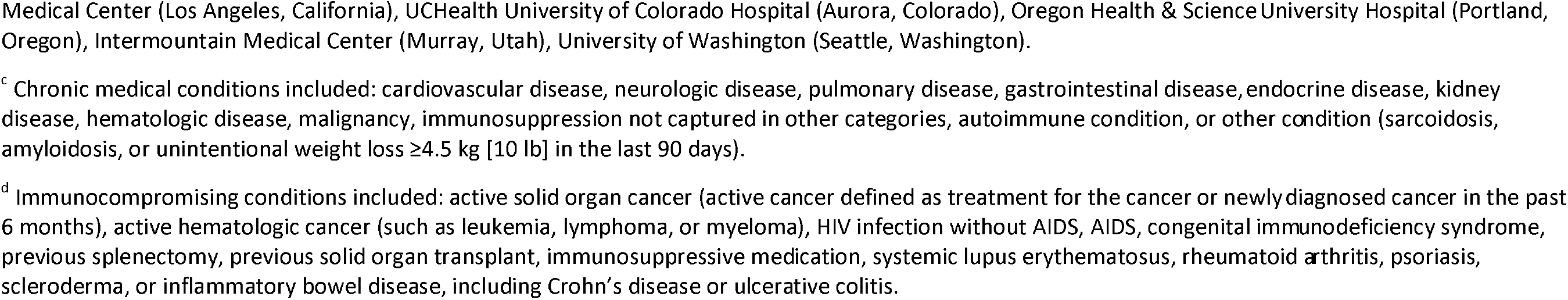
Baseline characteristics by vaccination and COVID-19 status—21 hospitals in 18 US states, December 26, 2021–April 30, 2022 (SARS-CoV-2 Omicron variant period)

Of the 1,572 case-patients, 671 (43%) were unvaccinated, 654 (42%) completed a primary series alone, and 247 (16%) completed a primary series plus one booster dose. Of the 1,609 control-patients, 386 (24%) were unvaccinated, 672 (42%) completed a primary series alone, and 551 (34%) received a primary series plus one booster dose. Unvaccinated patients were younger (median 58 years [IQR 44– 69]) than those who completed a primary series alone (median 65 years [IQR 54–74]), or a primary series plus one booster (median 68 years [IQR 59–78]) (p<0.001). Unvaccinated patients were less likely to have had ≥1 hospital admission in the past year than those who were vaccinated with a primary series plus one booster dose or primary series alone (p<0.001) (**Table 1**).

Viral whole genome sequencing was performed for 648 COVID-19 case-patients, and a SARS-CoV-2 variant was successfully identified in 508 (78%). Among those with a variant identified, 444 (87%) were Omicron, and 64 (13%) were Delta. Patients with the Delta variant were excluded from analyses. Of the 444 sequenced Omicron cases, 396 (89%) were BA.1 lineage and 48 (11%) were BA.2 lineage. BA.2 became the predominant lineage during the week starting March 27, 2022 (**Figure 1**).

**Figure 1.**
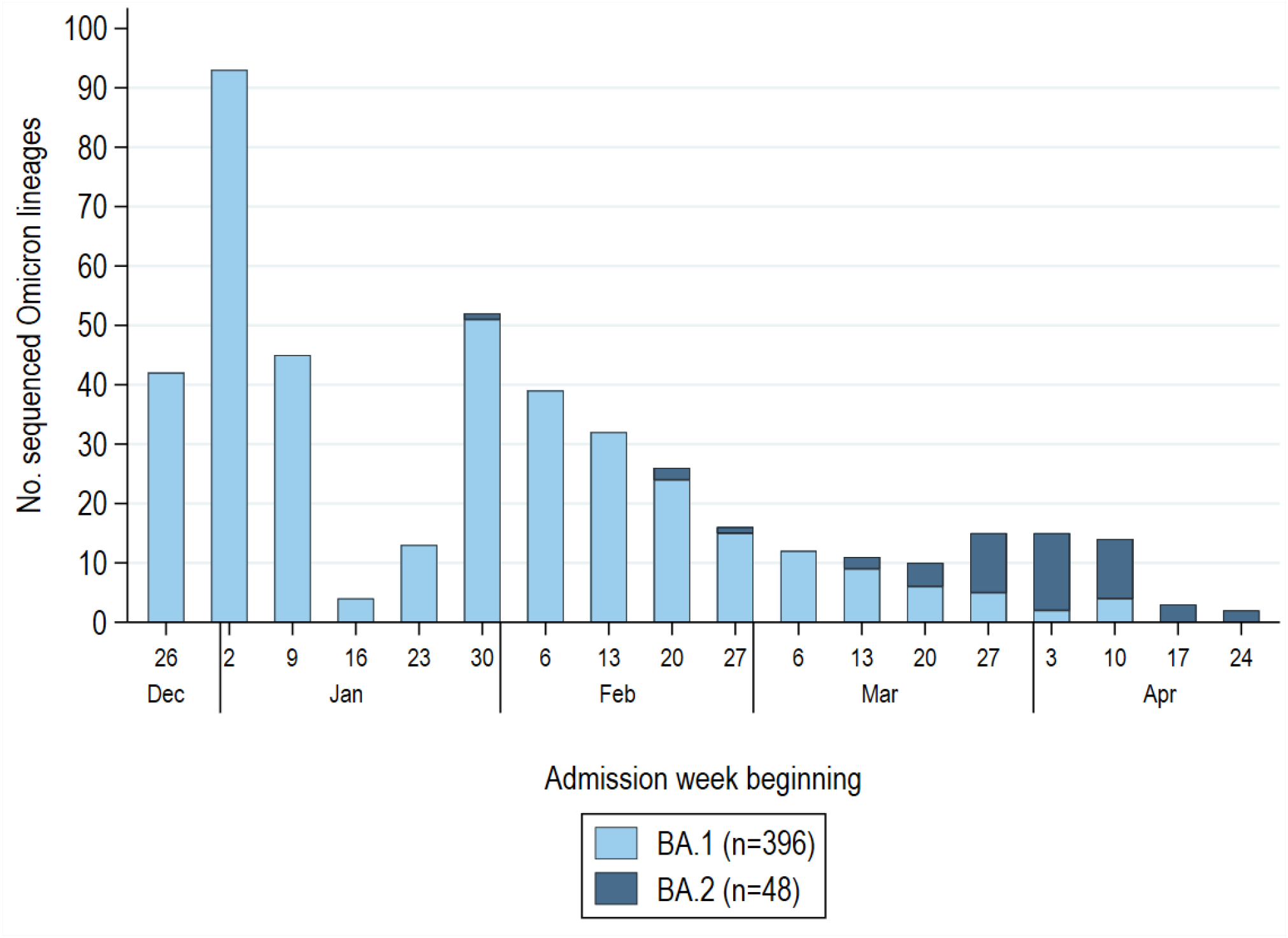
SARS-CoV-2 Omicron lineage by admission week among 1,572 COVID-19 case-patients enrolled during December 26, 2021–April 30, 2022 (with a pause in enrollment January 25–31, 2022). Sequenced Omicron variants were grouped into BA.1 (B.1.1.529, BA.1, BA.1.1, BA.1.15, BA.1.17) and BA.2 (BA.2, BA.2.1, BA.2.3) lineages. Non-Omicron case-patients (Delta variant, B.1.1.519) confirmed through sequencing were excluded from the analysis (n=64) and not displayed in this figure. Of 444 patients with a sequence-confirmed Omicron variant infection, 396 (89%) had BA.1 and 48 (11%) had BA.2. Low sequencing totals in late January reflect a pause in IVY network enrollment during January 25–31, 2022 during a protocol update.

Among vaccinated patients, most received a homologous mRNA primary series (90%), with lower proportions of patients receiving Ad26.COV2 (9%) or a heterologous mRNA (1%) primary series **(Figure 2)**. For booster dose product received, 97% of patients received an mRNA vaccine, with only 3% receiving Ad26.COV2. Most mixing of vaccine products occurred after receipt of one dose of Ad26.COV2 or after two doses of the same mRNA product.

**Figure 2.**
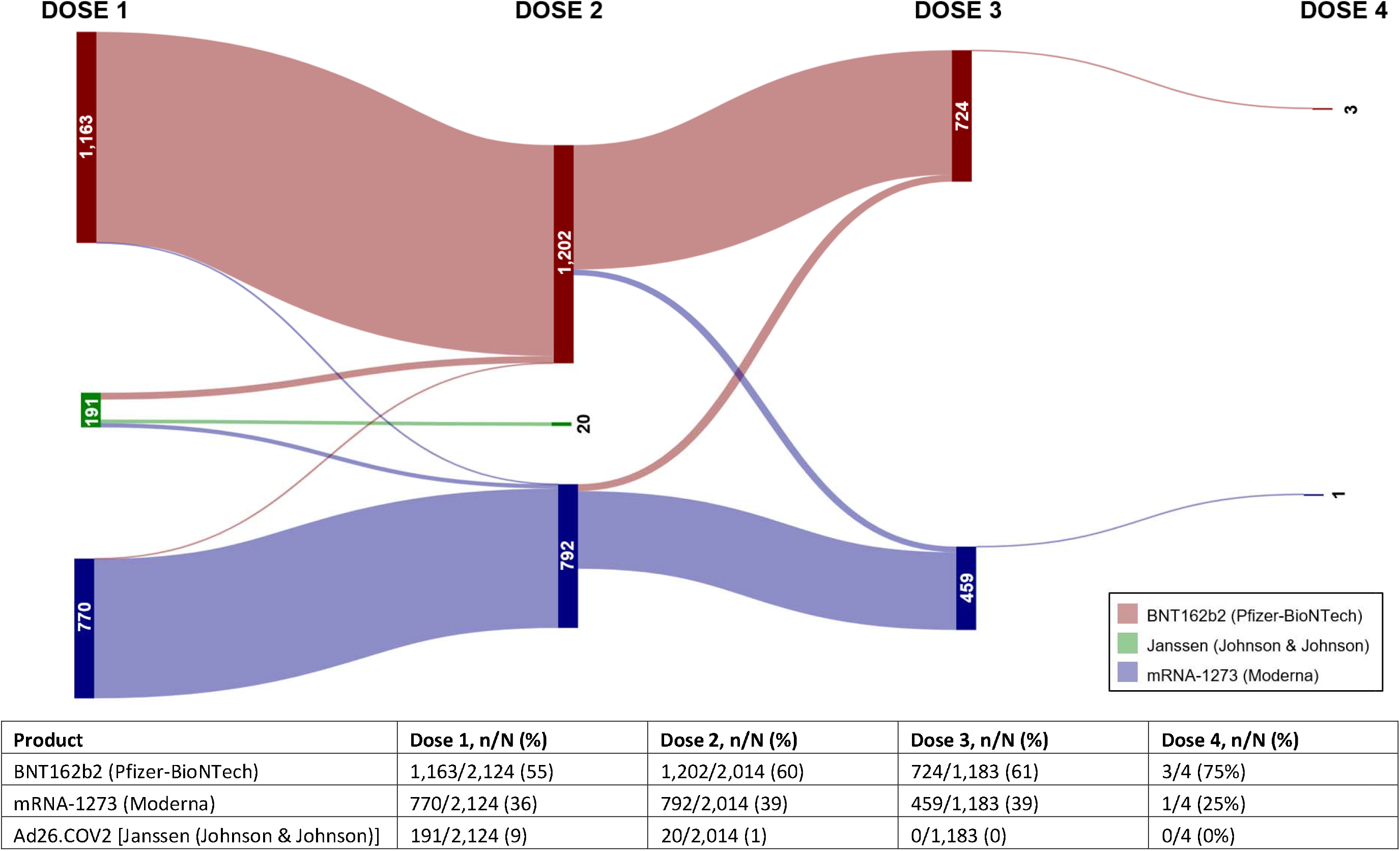
Pattern of COVID-19 vaccine products received across doses. The figure includes both patients hospitalized with COVID-19 (cases) and patients hospitalized with an acute respiratory illness without COVID-19 (controls) during December 26, 2021–April 30, 2022 (the SARS-CoV-2 Omicron variant period).

### VE against COVID-19 hospitalization

When considering all vaccine products (BNT162b2, mRNA-1273, and AD26.COV2), among immunocompetent patients, VE against COVID-19 hospitalization was higher for a primary series plus a booster (77%; 95% CI: 71–82) than a primary series alone (44%; 95% CI: 31–54) (p<0.001) **(Figure 3)**. Among immunocompetent patients vaccinated with either BNT162b2 (Pfizer) or mRNA-1273 (Moderna), those who received a homologous booster dose (i.e., a third dose of the same vaccine product) had significantly higher VE against COVID-19 hospitalization than those who received only a primary series. For BNT162b2, VE for a primary series plus a booster was 80% (95% CI: 73–85%) and for a primary series alone was 46% (95% CI: 30–58%) (p<0.001). For mRNA-1273, VE for a primary series plus a booster was 77% (95% CI: 67–83) and for a primary series alone was 47% (95% CI: 30–60) (p<0.001). While the sample size of patients who received a homologous booster dose with Ad26.COV2 (Johnson & Johnson) was small (n=20), the addition of a booster dose did not appear to increase VE compared to an AD26.COV2 single dose primary series. VE against COVID-19 hospitalization for two doses of Ad26.COV2 (i.e., a booster dose following a primary Ad26.COV2 dose) was 30% (95% CI: −85–74%) and for a single dose primary series alone was 41% (95% CI: 9–62%) (p=0.665). Among heterologous booster product recipients (those who received a different product for the primary series and booster dose), VE for mixed mRNA vaccine doses (i.e., any combination of BNT162b2 and mRNA-1273) was 70% (95% CI: 34– 86), while VE for a primary series Ad26.COV2 plus one mRNA booster was 64% (95% CI: 35–80).

**Figure 3.**
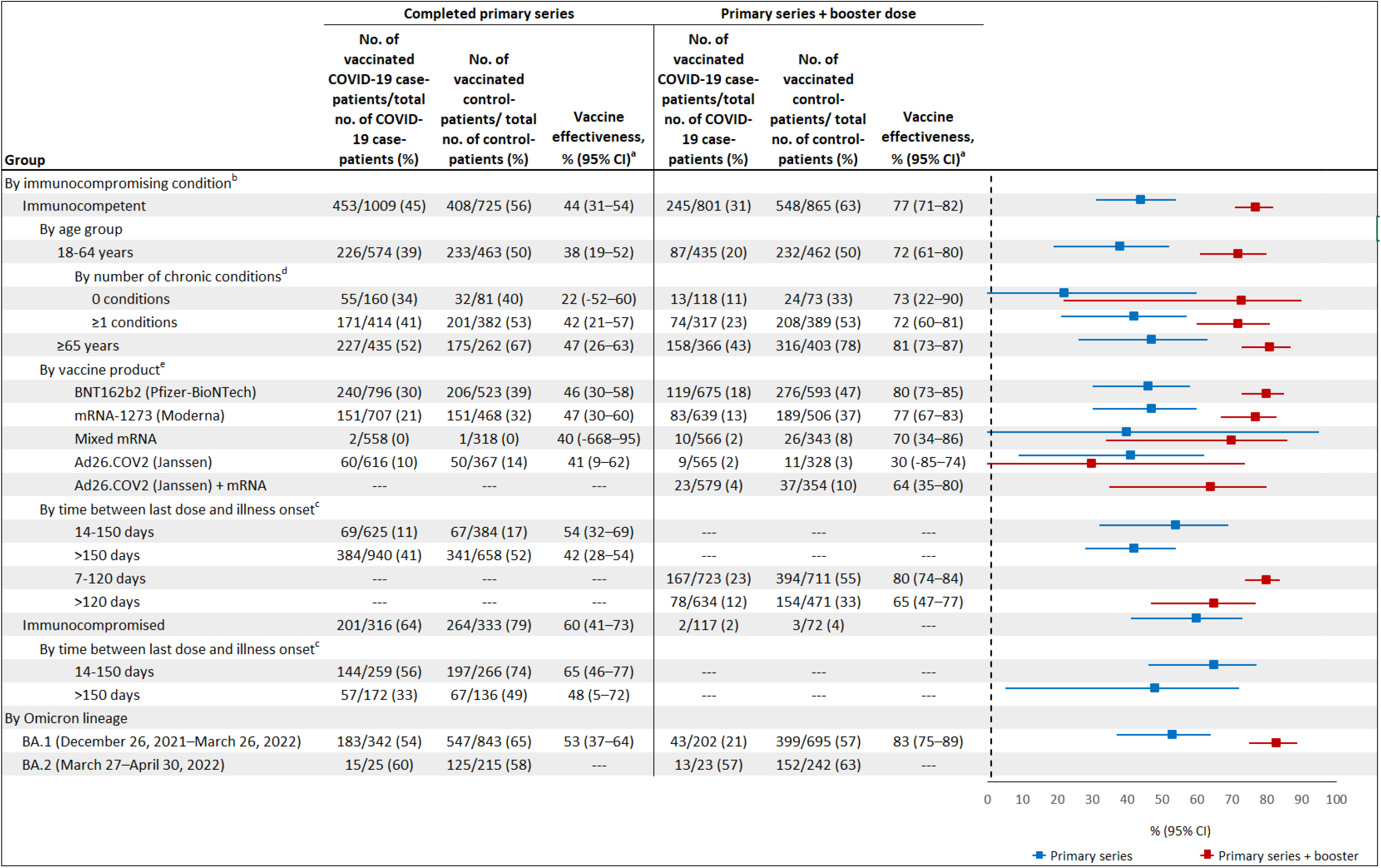
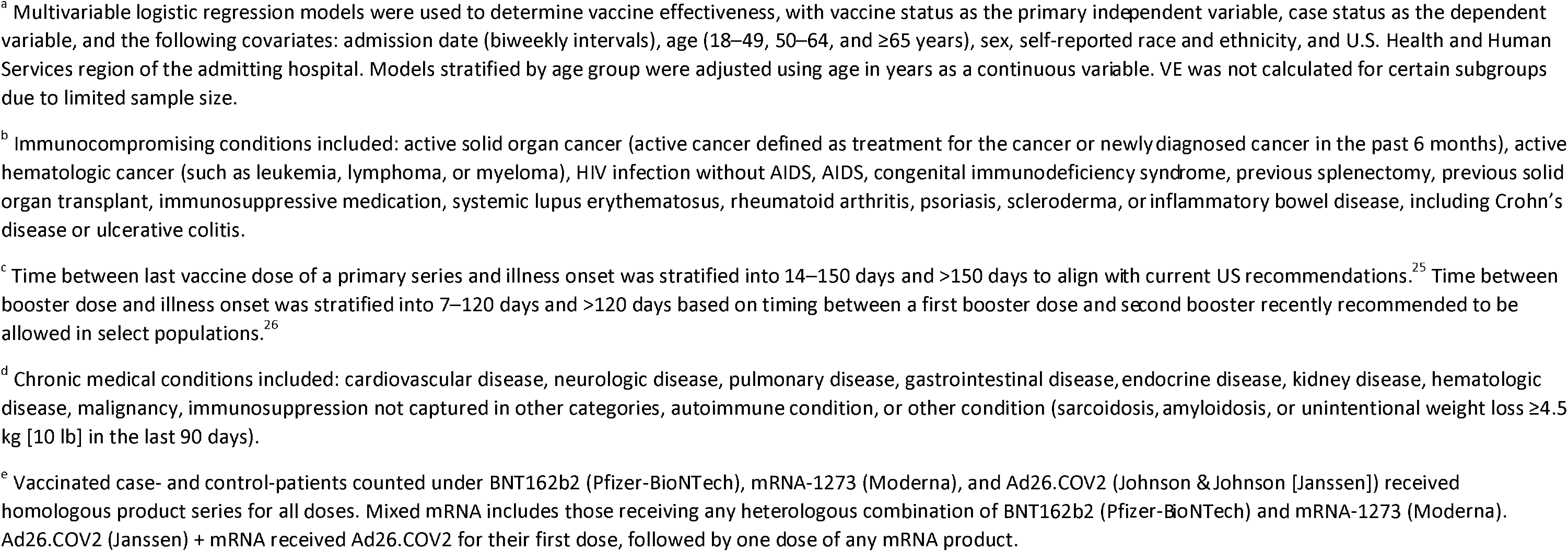
Vaccine effectiveness (VE) for prevention of COVID-19 hospitalization in the US during a period of SARS-CoV-2 Omicron variant predominance, December 26, 2021 through April 30, 2022. Vaccine effectiveness for a primary vaccine series plus one booster dose is displayed in red while vaccine effectiveness for a primary series alone is displayed in blue.

VE for immunocompromised patients completing a primary series (three doses of an mRNA vaccine) was 60% (95% CI: 41–73). VE could not be estimated for a primary series plus one booster in immunocompromised patients due to a small number of participants (n=5) who received a first booster dose (i.e., a fourth mRNA vaccine dose).

In analyses of VE against COVID-19 hospitalization stratified by time since the last vaccine dose, point estimates consistently showed a waning of protection at longer time points for both a primary series plus booster dose and a primary series alone. VE after a primary series for immunocompetent patients at 14–150 days (median 109 days) since the last vaccine dose was 54% (95% CI: 32–69), and at >150 days (median 279 days) was 42% (95% CI: 28–54%). VE after a booster dose for immunocompetent patients at 7–120 days (median 69 days) following the booster dose was 80% (95%: 74–84%) and at >120 days (median 147 days) was 65% (95% CI: 47–77%). For immunocompromised patients, VE for a primary series at 14–150 days (median 91 days) was 65% (95% CI: 46–77%) and at >150 days (median 172 days) was 48% (95% CI: 5–72%).

### Clinical outcomes among unvaccinated and vaccinated patients hospitalized with COVID-19

Clinical outcomes by day 28 of admission were available for the 1,361 COVID-19 case-patients admitted to hospital between December 26, 2021 and April 2, 2022; 1,117 (82%) of these patients were discharged from the hospital prior to day 28, 127 (9%) remained hospitalized at day 28, and 117 (9%) died in the hospital before day 28 (**Table 2**). Intensive care unit admission was more common among unvaccinated patients (34%) than vaccinated patients (26%, including 26% among those who completed a primary series alone and 26% among those who completed a primary series plus booster) (p=0.007). The composite critical COVID-19 outcome of death or invasive mechanical ventilation was more common among unvaccinated patients (23%) than vaccinated patients (15%, including 16% among those who completed a primary series alone and 14% among those who completed a primary series plus one booster dose) (p=0.002). Among 1,257 COVID-19 case-patients with data available on in-hospital treatments, use of one or more treatment indicated for severe COVID-19 (corticosteroids, tocilizumab, or baricitinib) was more common among unvaccinated patients (72%) than vaccinated patients (59%, including 60% who completed a primary series alone and 55% who completed a primary series plus booster) (p<0.001).

**Table 2.** In-hospital clinical outcomes and treatments for adults hospitalized with COVID-19 during December 26, 2021–April 2, 2022 (the SARS-CoV-2 Omicron variant period), by vaccination status.

| Outcome | Total | Unvaccinated | Primary vaccine series alone | Primary vaccine series plus booster | P-value<sup>a</sup> |
| --- | --- | --- | --- | --- | --- |
| Status at 28 days following admission, No. / Total (%) |  |  |  |  | 0.032 |
| Remained hospitalized | 127/1,361 (9%) | 65/621 (10%) | 55/570 (10%) | 7/170 (4%) |  |
| Discharged | 1,117/1,361 (82%) | 493/621 (79%) | 475/570 (83%) | 149/170 (88%) |  |
| Died | 117/1,361 (9%) | 63/621 (10%) | 40/570 (7%) | 14/170 (8%) |  |
| Admitted to intensive care unit, No. / Total (%) | 406/1,359 (30%) | 212/621 (34%) | 149/568 (26%) | 45/170 (26%) | 0.007 |
| Any oxygen support, No. / Total (%) | 1,022/1,361 (75%) | 488/621 (79%) | 410/570 (72%) | 124/170 (73%) | 0.023 |
| HFNC | 320/1,361 (24%) | 176/621 (28%) | 120/570 (21%) | 24/170 (14%) | <0.001 |
| NIPPV | 179/1,361 (13%) | 94/621 (15%) | 69/570 (12%) | 16/170 (9%) | 0.092 |
| IMV | 208/1,361 (15%) | 125/621 (20%) | 66/570 (12%) | 17/170 (10%) | <0.001 |
| ECMO | 18/1,361 (1%) | 15/621 (2%) | 3/570 (1%) | 0/170 (0%) | 0.005 |
| Vasopressors, no. (%) | 208/1,361 (15%) | 114/621 (18%) | 77/570 (14%) | 17/170 (10%) | 0.008 |
| New renal replacement therapy, no. (%) | 56/1,361 (4%) | 31/621 (5%) | 22/570 (4%) | 3/170 (2%) | 0.16 |
| Venous thromboembolic event, No. (%)<sup>b</sup> | 108/1,361 (8%) | 60/621 (10%) | 38/570 (7%) | 10/170 (6%) | 0.092 |
| Stroke, No. (%) | 20/1,361 (1%) | 11/621 (2%) | 6/570 (1%) | 3/170 (2%) | 0.56 |
| Myocardial infarction, No. (%) | 35/1,361 (3%) | 15/621 (2%) | 13/570 (2%) | 7/170 (4%) | 0.39 |
| Composite of death or IMV, no. (%) | 255/1,361 (19%) | 141/621 (23%) | 90/570 (16%) | 24/170 (14%) | 0.002 |
| Any of the following therapeutics used for severe COVID-19, No. / Total (%) | 812/1,257 (65%) | 404/561 (72%) | 318/533 (60%) | 90/163 (55%) | <0.001 |
| Corticosteroids | 808/1,257 (64%) | 403/561 (72%) | 316/533 (59%) | 89/163 (55%) | <0.001 |
| Tocilizumab | 48/1,257 (4%) | 28/561 (5%) | 16/533 (3%) | 4/163 (2%) | 0.14 |
| Baricitinib | 90/1,257 (7%) | 64/561 (11%) | 26/533 (5%) | 0/163 (0%) | <0.001 |
| Anti-SARS-CoV-2 Monoclonal Antibodies | 29/1,257 (2%) | 11/561 (2%) | 16/533 (3%) | 2/163 (1%) | 0.32 |

Abbreviations: ECMO = extracorporeal membrane oxygenation; HFNC = high-flow nasal cannula; ICU = intensive care unit; IMV = invasive mechanical ventilation; NIPPV = nasal intermittent positive-pressure ventilation
<sup>a</sup> Statistical differences were evaluated using Pearson's chi-squared test for binary or categorical variables. Statistical significance was determined using a p-value threshold of 0.05. <sup>b</sup> Deep vein thrombosis and/or pulmonary embolism

### Pre-booster and post-booster serum antibody results

Paired serum samples were collected before and after a COVID-19 booster dose in 63 healthy volunteers, including 33 with a BNT162b2 booster dose after a BNT162b2 primary series, 16 with an mRNA-1273 booster dose following an mRNA-1273 primary series, 8 with an Ad26.COV2 booster dose after an Ad26.COV2 primary series dose and 6 with one mRNA booster dose after an Ad26.COV2 primary series dose **(Table S5)**. Anti-RBD and anti-S antibody concentrations were higher after compared to before the booster dose for all vaccines **(Figure 4; Table S6)**. Post-booster anti-RBD and anti-S antibody concentrations were higher for participants who received three BNT162b2 doses and three mRNA-1273 doses compared to two Ad26.COV2 doses.

**Figure 4.**
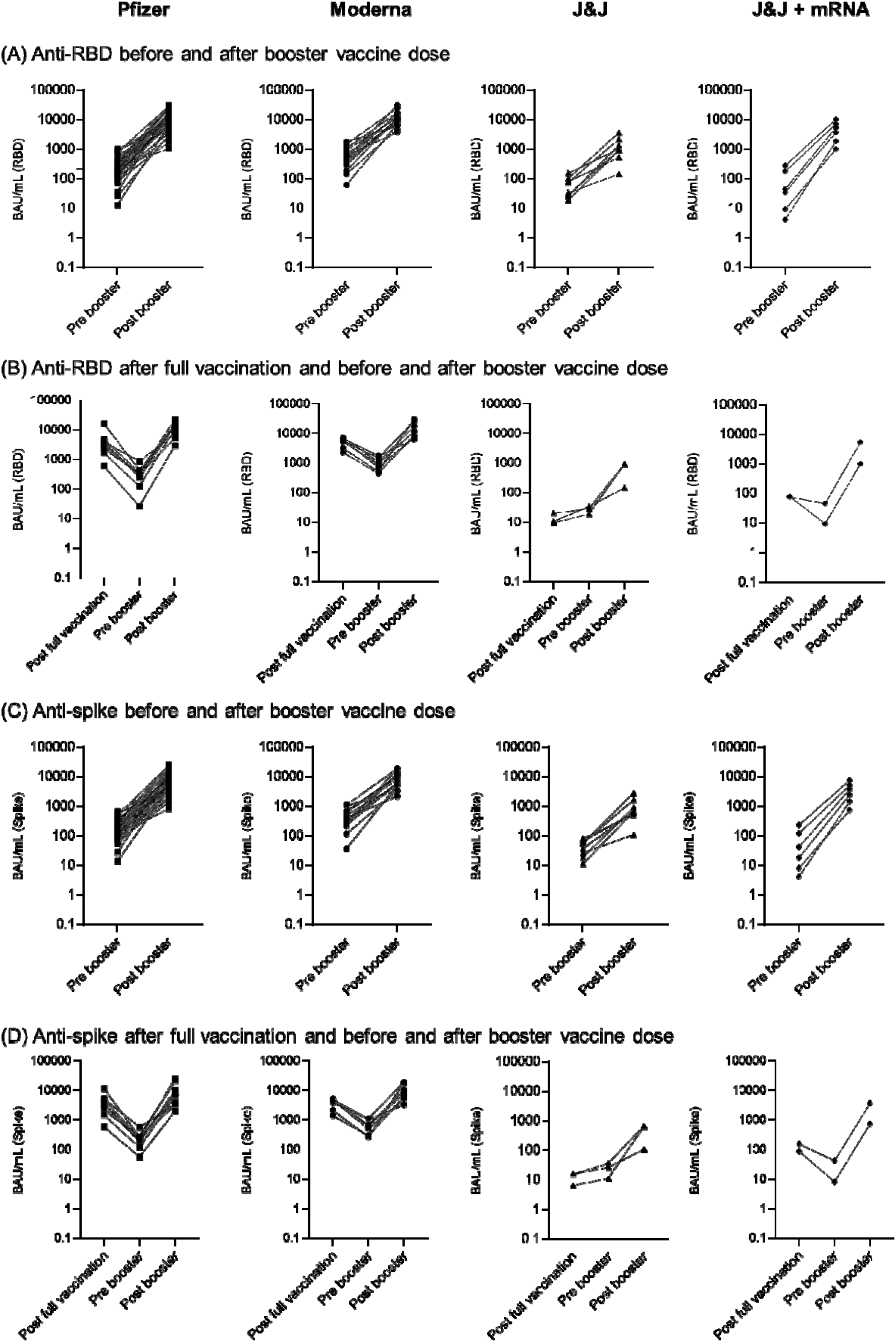
Spaghetti plots of serum antibody concentrations to the SARS-CoV-2 receptor binding domain (anti-RBD) (panel A) and spike protein (anti-spike) (panel C) among healthy adult volunteers before and 2–6 weeks after receiving COVID-19 booster doses, October 5, 2021–January 28, 2022. Targets of the antibody responses were based on proteins from the USA-WA1/2020 strain. Antibody concentrations 2– 6 weeks after a primary series is also displayed for the subset of participants with anti-RBD (panel B) and anti-spike (panel D) measurements available from prior participation in the program. Each participant is represented with a single line connecting the antibody concentration at each time point. The Pfizer-BioNTech group included 33 participants who received three doses of BNT162b2, including 9 who had antibody measurements after the second dose. The Moderna group included 16 participants who received three mRNA-1273 doses, including 7 who had antibody measurements after the second dose. The Johnson and Johnson (J&J) group included 8 participants who received two doses of Ad26.COV2, including 3 who had antibody measurements after the first dose. The J&J + mRNA group included 6 participants who received an mRNA vaccine (BNT162b2 or mRNA-1273) booster dose after a single Ad26.COV2 primary series dose, including 2 who had antibody measurements after the initial Ad26.COV2 dose. The data accompanying this figure are shown in Table S6.

## DISCUSSION

### Principal Findings

In this analysis of hospitalized adults at 21 hospitals in the US during an Omicron predominant period from December 26, 2021–April 30, 2022, receipt of a COVID-19 primary vaccine series plus a booster dose provided greater protection against COVID-19 hospitalization than receipt of a primary series alone. This added protection of a booster dose was seen across all age groups and for all three vaccine products used in the US. mRNA vaccines (BNT162b2 and mRNA-1273) provided greater protection than the Ad26.COV2 vaccine, both when considering a primary series alone and boosted vaccine regimens. These clinical vaccine effectiveness findings were supported by serology results demonstrating substantially higher anti-SARS-CoV-2 antibody titers following booster vaccine doses. These antibody and VE data support US recommendations that all eligible patients receive a booster dose to protect against severe disease from the Omicron variant.

### Comparison with other studies and policy implications

COVID-19 vaccines have been authorized in the US for over a year, and as of June 2022, an estimated 77% of the adult population has received a primary series, of whom nearly half of those eligible to receive a first booster dose had not yet received it.^3^ Modest waning in protection of a primary COVID-19 vaccine series was initially demonstrated during the Delta variant period.^29–31^ Our findings support the concept of waning protection for primary vaccine series for the Omicron variant as well, with a decline in protection for a primary series against COVID-19 hospitalization during the Omicron period greater than 5 months after a primary series. These data add to the growing evidence of waning effectiveness of a primary series.^32,33^ Despite known immune evasion features of the Omicron variant for vaccines currently available, our results show that VE substantially increased following a booster dose, consistent with results from other studies.^7,8^ Data from immunological studies suggest that a broadening of the immune response occurs after a booster dose, including an increase in, and adaptation of, anti-receptor binding domain-specific memory B cells, providing biologic plausibility for increased VE after a booster dose even with the highly divergent Omicron variant.^25,34–36^ Our data from healthy adult volunteers suggested significant increases in anti-RBD and anti-spike antibody levels 2-6 weeks after receiving one booster dose, consistent with observed differences in VE between those who received a primary series only versus those who received one booster dose. Understanding the extent to which VE against severe Omicron disease wanes following booster doses will require the accumulation of more data over time. While some early evidence from the US, United Kingdom, and Israel have shown some decrease in effectiveness of mRNA vaccines >120 days from a first booster dose^10,37,38^, we found only a modest decline in VE following a booster dose in this study (from 80% at 7–120 days to 65% at >120 days).

We also observed that vaccinated patients hospitalized with COVID-19 experienced less severe outcomes than unvaccinated patients hospitalized with COVID-19, suggesting that COVID-19 vaccines result in disease attenuation and providing further rationale for COVID-19 vaccination.^30,31^ In this analysis, while patients who received boosted vaccine regimens were less likely to be hospitalized with COVID-19 than those who received a primary series alone, in-hospital outcomes were similar between those who received boosted regimens and a primary series alone. Additional study will be required to understand if booster doses lead to incremental disease attenuation beyond a primary series among vaccine breakthrough cases.

Our findings also suggest lower effectiveness for the adenovirus vector vaccine (Ad26.COV2) both as a primary vaccine series and as a booster dose compared to mRNA vaccines. Although mechanisms of protection against SARS-CoV-2 have not yet been established, binding antibodies correlate with protection against SARS-CoV-2, allowing for contextualization of findings from observational data.^27,39,40^ In this analysis, the persistently low anti-spike and anti-RBD levels post-primary series dose and before a first booster dose for recipients of Ad26.COV2 compared with recipients of mRNA vaccines provide complementary understanding of the reduced effectiveness of this vaccine relative to mRNA vaccines. These findings support preferential use of mRNA vaccines both for a primary series and as a booster dose, regardless of which vaccine product was used for the primary series.^25,26^

### Strengths and limitations

Important strengths of this study included enrollment of patients with active COVID-19 symptoms ascertained at the bedside in real-time and laboratory confirmed SARS-CoV-2 infection; these methods facilitated calculations of vaccine effectiveness to prevent symptomatic, laboratory confirmed COVID-19 hospitalization. Additionally, ascertainment of vaccination status was robust, with patient interviews combined with systematic searches of medical records and vaccines registries, which enabled precise classification of each patient’s vaccination status, including the receipt of booster doses. Furthermore, respiratory samples with SARS-CoV-2 detection underwent viral sequencing, which enabled exclusion of patients identified to have infections with a variant other than Omicron.

This analysis also had limitations. First, although several relevant confounders were controlled in VE models, residual or unmeasured confounding is possible. In particular, increases in the proportion of individuals with prior SARS-CoV-2 infection, which is associated with a degree of protection against reinfection, may have led lower VE observed in this study compared with studies from earlier in the pandemic when fewer control patients had prior natural infection.^41-43^ Second, although binding antibodies likely correlate with protection against SARS-CoV-2 infection, other immune responses including neutralizing antibodies and cellular immunity were not evaluated. Third, patients included in the antibody analysis were healthy adult volunteers and may not fully represent the clinical and demographic characteristics of those included in VE analyses. Fourth, modest sample size limited precision of some estimates, including waning of VE >120 days after a booster dose and effectiveness of Ad26.COV2, and prevented estimates for effectiveness of booster doses in immunocompromised patients. Fifth, our study was restricted to analysis of first booster doses due to second booster doses only emerging in select populations toward the end of the study period and low uptake of second boosters in the study population. Future studies on second boosters will be important.

## CONCLUSION

Vaccination with a primary COVID-19 vaccine series plus a booster dose was significantly more effective than a primary series alone in preventing COVID-19 associated hospitalization during the Omicron-variant period in the US. Additionally, mRNA vaccines were more effective than the Ad26.COV2 vaccine, both as a primary series and as booster doses. Serology results support these findings by showing substantial increases in anti-SARS-CoV-2 antibody titers following booster vaccine doses, particularly with mRNA vaccines. These findings support recommendations for all eligible people aged ≥18 years to receive a booster dose of an mRNA vaccine.

## Data Availability

No additional data are available

## NOTES

### Contributions

Guarantors of this work include Dr. Self (protocol and data integrity), Ms. Adams (statistical analysis), Dr. Lauring (viral sequencing laboratory methods), Dr. Chappell (RT-PCR laboratory methods), Dr. Thornburg (antibody measurement methods). Contributions of each author include the following. Responsibility for decision to submit the manuscript: Self. Composed the initial manuscript draft: Adams, Rhoads, Surie, Tenforde, Self (the authors alone wrote the manuscript without outside assistance). Conceptualization of study methods: Adams, Rhoads, Surie, Gaglani, Ginde, McNeal, Ghamande, Huynh, Talbot, Casey, Mohr, Zepeski, Shapiro, Gibbs, Files, Hicks, Hager, Ali, Prekker, Frosch, Exline, Gong, Mohamed, Johnson, Srinivasan, Steingrub, Peltan, Brown, Martin, Monto, Lauring, Khan, Hough, Busse, ten Lohuis, Duggal, Wilson, Gordon, Qadir, Chang, Mallow, Rivas, Babcock, Kwon, Chappell, Halasa, Grijalva, Rice, Stubblefield, Baughman, Lindsell, Hart, Lester, Thornburg, Park, McMorrow, Patel, Tenforde, Self. Statistical analysis and data management: Adams, Tenforde, Lindsell, Hart, Zhu. Funding acquisition: Self. Critical review of the manuscript for important intellectual content: Adams, Rhoads, Surie, Gaglani, Ginde, McNeal, Ghamande, Huynh, Talbot, Casey, Mohr, Zepeski, Shapiro, Gibbs, Files, Hicks, Hager, Ali, Prekker, Frosch, Exline, Gong, Mohamed, Johnson, Srinivasan, Steingrub, Peltan, Brown, Martin, Monto, Lauring, Khan, Hough, Busse, ten Lohuis, Duggal, Wilson, Gordon, Qadir, Chang, Mallow, Rivas, Babcock, Kwon, Chappell, Halasa, Grijalva, Rice, Stubblefield, Baughman, Lindsell, Hart, Lester, Thornburg, Park, McMorrow, Patel, Tenforde, Self. The corresponding author (Dr. Self) attests that all listed authors meet authorship criteria and that no others meeting the criteria have been omitted.

### Patient Consent

not applicable.

### Ethical Approval

This program was approved as a public health surveillance activity with waiver of informed consent by institutional review boards at the US Centers for Disease Control and Prevention (CDC), the program’s coordinating center at Vanderbilt University Medical Center and each participating site.

### Data Sharing

No additional data are available.

### Transparency

Dr. Self affirms that the manuscript is an honest, accurate, and transparent account of the study being reported; that no important aspects of the study have been omitted; and that any discrepancies from the study as planned have been explained. Dr. Self led the development of the study protocol and participant enrollment. Ms. Adams led statistical analysis. Dr. Lauring led viral sequencing work. Dr. Chappell led RT-PCR work. Dr. Self takes responsibility for the work overall.

### Dissemination to participants and related patient and public communities

Results will be disseminated to relevant communities via public health announcements from the US Centers for Disease Control and Prevention, via press releases in the lay press, and public presentations by the investigators.

