## supplement for "Vaccine Effectiveness of Primary Series and Booster Doses against Omicron Variant COVID-19-Associated Hospitalization in the United States"

IVY

### Supplementary Materials

**Vaccine Effectiveness of Primary Series and Booster Doses against Omicron Variant COVID-19-Associated Hospitalization in the United States**

The IVY Network

Corresponding Author:
Wesley H. Self, MD, MPH; Vanderbilt University Medical Center; Nashville, Tennessee USA.
.

INFLUENZA AND OTHER VIRUSES IN THE ACUTELY ILL

**Supplementary Appendix A: Investigators and Collaborators**

Investigators and collaborators in the Influenza and Other Viruses in the Acutely Ill (IVY) Network

**Baylor, Scott and White, Temple, Texas**
Manjusha Gaglani, Tresa McNeal, Shekhar Ghamande, Nicole Calhoun, Kempapura Murthy, Judy Herrick, Amanda McKillop, Eric Hoffman, Martha Zayed, Michael Smith

**Baystate Medical Center, Springfield, Massachusetts**
Jay Steingrub, Lori-Ann Kozikowski, Lesley De Souza, Scott Ouellette

**Beth Israel Medical Center, Boston Massachusetts**
Nathan I. Shapiro, Michael Bolstad, Brianna Coviello, Robert Ciottone, Arnaldo Devilla, Ana Grafals, Conor Higgins, Carlo Ottanelli, Kimberly Redman, Douglas Scaffidi, Alexander Weingart

**Centers for Disease Control and Prevention (CDC), Atlanta, Georgia**
Manish Patel, Mark Tenforde, Nathaniel Lewis, Samantha Olson, Meagan Stephenson, Katherine Adams, Diya Surie, Meredith McMorrow, Maraia Tremarelli, Caitlin Turbyfill

**Cleveland Clinic, Cleveland, Ohio**
Omar Mehkri, Megan Mitchell, Zachary Griffith, Connery Brennan, Kiran Ashok, Bryan Poynter, Abhijit Duggal

**Emory University, Atlanta, Georgia**
Laurence Busse, Caitlin ten Lohuis, Nicholas Stanley, Sophia Zhang

**Hennepin County Medical Center, Minneapolis, Minnesota**
Matthew Prekker, Heidi Erickson, Anne Frosch, Audrey Hendrickson, Sean Caspers, Walker Tordsen, Olivia Kaus, Tyler Scharber

**Intermountain Medical Center, Murray, Utah**
Ithan Peltan, Samuel Brown, Jenna Lumpkin, Cassie Smith, Hunter Marshall

**Johns Hopkins University, Baltimore, Maryland**
David N, Hager, Arber Shehu, Harith Ali, Richard E. Rothman

**Montefiore Medical Center, Bronx, New York**
Michelle Gong, Amira Mohamed, Rahul Nair, Jen-Ting (Tina) Chen

**Ohio State Medical Center, Columbus, Ohio**
Matthew Exline, Sarah Karow, Emily Robart, Paulo Nunes Maldonado, Maryiam Khan, Preston So, Madison So, Elizabeth Schwartz, Mena Botros

**Oregon Health and Sciences University, Portland, Oregon**
Akram Khan, Catherine L. Hough, Haeun Jung, Jesus Martinez, Andrea Luong, Bao Huynh, Habiba Ibrahim, Cynthia Villanueva-Vargas, Juliana Villanueva-Vargas, Suha Quadri

**Stanford University, Stanford, California**
Jennifer G. Wilson, Alexandra June Gordon, Joe Levitt, Cynthia Perez, Anita Visweswaran, Jonasel Roque

**University of California-Los Angeles, Los Angeles, California**
Nida Qadir, Steven Chang, Trevor Frankel, Omai Garner, Sukantha Chandrasekaran

**University of Colorado, Aurora, Colorado**
Adit Ginde, David Douin, Kelly Jensen, David Huynh, Aimee Steinwand, Cori Withers

**University of Iowa, Iowa City, Iowa**
Nicholas Mohr, Anne Zepeski, Paul Nassar, Shannon Landers, Karin Nielsen, Noble Briggs, Cathy Fairfield

**University of Miami, Miami, Florida**
Chris Mallow, Hayley Gershengorn, Carolina Rivas

**University of Michigan, Ann Arbor, Michigan**
Emily Martin, Arnold Monto, Adam Lauring, EJ McSpadden, Rachel Truscon, Anne Kaniclides, Lara Thomas, Ramsay Bielak, Weronika Damek Valvano, Rebecca Fong, William J. Fitzsimmons, Christopher Blair, Julie Gilbert, Leigh Baker

**University of Washington, Seattle, Washington**
Nicholas Johnson, Vasisht Srinivasan, Christine D. Crider, Kyle A. Steinbock, Thomas C. Paulsen, Layla A. Anderson

**Vanderbilt University Medical Center, Nashville, Tennessee**
Wesley H. Self, H. Keipp Talbot, Chris Lindsell, Carlos Grijalva, Ian Jones, Natasha Halasa, James Chappell, Kelsey Womack, Jillian Rhoads, Adrienne Baughman, Christy Kampe, Jakea Johnson, Jake Sturgill, Kim Hart, Robert McClellan, Todd Rice, Jonathan Casey, William B. Stubblefield, Yuwei Zhu, Laura L. Short, Lauren J. Ezzell, Margaret E. Whitsett, Rendie E. McHenry, Samarian J. Hargrave, Marcia Blair, Jennifer L. Luther, Claudia Guevara Pulido, Bryan P. M. Peterson

**Wake Forest University, Winston-Salem, North Carolina**
D. Clark Files, Kevin Gibbs, Mary LaRose, Leigha Landreth, Madeline Hicks, Lisa Parks

**Washington University, St. Louis, Missouri**
Hilary Babcock, Jennie Kwon, Jahnavi Bongu, David McDonald, Candice Cass, Sondra Seiler, David Park, Tiffany Hink, Meghan Wallace, Carey-Ann Burnham, Olivia G. Arter

Appendix B: Supplementary Tables

Table S1. IVY Network COVID-19 vaccine effectiveness publications.
The IVY Network publishes vaccine effectiveness (VE) estimates iteratively, with later publications adding sample size and focusing on specific questions that were not addressed in earlier publications. Some participants are included in multiple publications. The current manuscript includes participants enrolled during December 26, 2021–April 30, 2022 and focuses on vaccine effectiveness against the Omicron variant.

| Publication | Primary Question Addressed | Data cut (cohort initiated March 11, 2021) | Sample size | Primary finding |
|---|---|---|---|---|
| Clin Infect Dis 2021. PMID: 34358310. | COVID-19 mRNA VE against hospitalization during the early phase of the US COVID-19 program. | May 5, 2021 | 1,212 | mRNA VE against COVID-19 hospitalization among US adults = 87.1% (95% CI: 81–91) |
| MMWR 2021. PMID: 34437524 | COVID-19 mRNA VE against hospitalization beyond 12 weeks from full vaccination. | July 14, 2021 | 3,089 | mRNA VE against COVID-19 hospitalization from 13–24 weeks post-vaccination among US adults = 84% (95% CI: 77–90) |
| MMWR 2021. PMID: 34555004 | Comparative effectiveness of COVID-19 vaccines available in the United States for preventing COVID-19 hospitalizations among immunocompetent adults | August 15, 2021 | 3,689 | VE against COVID-19 hospitalizations varied by vaccine product among immunocompetent adults – Moderna: 93% (95% CI: 91–95); Pfizer BioNTech: 88% (95% CI: 85–91); Janssen: 71% (95% CI: 56–81). |
| JAMA 2021. PMID: 34734975 | Effectiveness of mRNA vaccines to prevent disease progression to critical illness or death among adults hospitalized with COVID-19. | August 15, 2021 | 4,513 | Prior vaccination was associated with a 67% (95% CI: 42–81) relative risk reduction lower risk of progression to invasive mechanical ventilation or death among adults hospitalized with COVID-19, suggesting vaccination attenuated disease severity. |
| MMWR 2022. PMID: 35085218 | Effectiveness of a Third Dose of Pfizer-BioNTech and Moderna Vaccines in Preventing COVID-19 Hospitalization Among Immunocompetent and Immunocompromised Adults - United States, August-December 2021 | August 19–December 15, 2021 | 2,952 | Three doses of mRNA COVID-19 vaccine provide greater protection against COVID-19 hospitalization than two doses. For immunocompetent adults, VE against hospitalization was 97% for three doses and 82% for two doses. For immunocompromised adults, VE against hospitalization was 88% for three doses and 69% for two doses. |

| Publication | Primary Question Addressed | Data cut (cohort initiated March 11, 2021) | Sample size | Primary finding |
|---|---|---|---|---|
| BMJ 2022; PMID: 35264324 | Clinical Severity of, and effectiveness of mRNA vaccines against, COVID-19 from Omicron, Delta, and Alpha SARS-CoV-2 Variants in the United States | January 14, 2022 | 11,690 | VE against COVID-19 hospitalization during the period that Delta variant dominated was 85% (95% CI: 83–87) for 2 mRNA vaccine doses and 94% (95% CI: 92–95) for 3 vaccine doses; for the early period that Omicron variant dominated VE was 65% (95% CI: 51–75) for 2 doses and 86% (95% CI: 77–91) for 3 doses. In-hospital COVID-19 severity was lower for Omicron than Delta. Prior vaccination was associated with lower in-hospital COVID-19 severity for Alpha, Delta, and Omicron variants. |
| MMWR 2022; PMID: 35324878 | mRNA vaccine effectiveness during the alpha and delta periods against invasive mechanical ventilation and death | January 24, 2022 | 7,544 | mRNA vaccine effectiveness to prevent death or invasive mechanical ventilation was 90% (95% CI = 88–91). |
| J Infect Dis 2022; PMID:35385875 | mRNA vaccine effectiveness against COVID-19 hospitalization among solid organ transplant recipients. | December 15, 2021 | 10,420 | VE against COVID-19 hospitalizations varied by host immune status with SOT recipients having lower VE for two vaccine doses (29%; 95% CI: -19–58) and three vaccine doses (77%, 95% CI: 48–90) than immunocompetent people. |

Table S2. Underlying medical conditions.

Conditions considered in this analysis to be immunocompromising are noted (*).

| |
|---|
| **Cardiovascular disease** |
| Heart failure |
| Peripheral vascular disease that limits mobility |
| Prior myocardial infarction |
| Cardiac arrhythmias including atrial fibrillation and ventricular arrhythmias |
| Valvular heart disease |
| Hypertension |
| **Neurologic disease** |
| Dementia |
| Prior stroke |
| Prior transient ischemic attack (TIA, “mini-stroke”) |
| Brain or spinal cord injury with loss of limb function |
| Cerebral palsy |
| Muscular dystrophy |
| Multiple sclerosis |
| Myasthenia gravis |
| Anterolateral sclerosis (ALS) |
| **Pulmonary disease** |
| Asthma |
| Chronic obstructive pulmonary disease |
| Cystic fibrosis |
| Pulmonary fibrosis |
| Pulmonary hypertension |
| Home oxygen use (except at night for sleep disorder) |
| Tracheostomy |
| Home non-invasive ventilation use (except at night for sleep disorder) |
| Home invasive ventilation use |
| **Gastrointestinal disease** |
| Feeding through a tube |
| Inflammatory bowel disease including Crohn's Disease or Ulcerative Colitis* |
| Cirrhosis (clinical diagnosis of cirrhosis) |
| Chronic liver disease without cirrhosis |
| Peptic ulcer disease |
| **Endocrine disease** |
| Diabetes mellitus without end organ damage |
| Diabetes mellitus with end organ damage |
| Adrenal insufficiency |
| Hypothyroidism |
| **Renal disease** |
| Chronic kidney disease without chronic renal replacement therapy |
| End stage renal disease on chronic renal replacement therapy (including hemodialysis or peritoneal dialysis) |
| Prior kidney transplant* |

| | |
|---|---|
| **Hematologic disease** | |
| | Sickle cell disease (all variants) |
| | Prior stem cell or bone marrow transplant* |
| | Coagulopathy or other bleeding disorder, such as hemophilia |
| | Chronic anemia |
| | Thalassemia |
| **Cancer/Malignancy** | |
| | Active solid organ cancer without metastases; active cancer defined as treatment for the cancer or newly diagnosed cancer in the past 6 months* |
| | Active solid organ cancer with metastases; active cancer defined as treatment for the cancer or newly diagnosed cancer in the past 6 months* |
| | Active hematologic cancer (such as leukemia/lymphoma/myeloma) or active cancer defined as treatment for the cancer or newly diagnosed cancer in the past 6 months* |
| **Other immunosuppression** | |
| | Human Immunodeficiency Virus (HIV) infection without AIDS* |
| | AIDS* |
| | Congenital immunodeficiency disorder* |
| | Prior splenectomy* |
| | Prior solid organ transplant* |
| | Immunosuppression medication* |
| | Other immunosuppression condition |
| **Autoimmune** | |
| | Systemic Lupus Erythematosus* |
| | Rheumatoid arthritis* |
| | Psoriasis* |
| | Scleroderma* |
| | Other autoimmune disease |
| **Other** | |
| | Unintentional weight loss of at least 10 pounds in past 90 days |
| | Sarcoidosis |
| | Amyloidosis |

Table S3. Classification of vaccination groups.

Definitions of vaccination groups included in this analysis —21 hospitals in 18 US states, December 26, 2021–April 30, 2022

| **Vaccination group** | **Immunosuppression status[a]** | **Doses received** | **Product** | | | | | | **Interval between last dose and illness onset** | **Patients included in this analysis (Total N=3,181)** |
|---|---|---|---|---|---|---|---|---|---|---|
| | | | **First dose** | **Second dose** | **Third dose** | **Fourth dose** | **Fifth dose** | **Sixth dose** | | |
| Unvaccinated | -- | 0 | -- | -- | -- | -- | -- | -- | -- | 1,057/3,181 (33%) |
| Partially vaccinated | -- | 1 | Any | -- | -- | -- | -- | -- | 0-13 days | Excluded |
| Partially vaccinated | Immunocompetent | 1 | mRNA[b] | -- | -- | -- | -- | -- | N/A | Excluded |
| Partially vaccinated | Immunocompetent | 2 | mRNA | mRNA | | | -- | -- | 0-13 days | Excluded |
| Partially vaccinated | Immunocompromised | 1 | mRNA | -- | -- | -- | -- | -- | N/A | Excluded |
| Partially vaccinated | Immunocompromised | 2 | mRNA | mRNA | -- | -- | -- | -- | N/A | Excluded |
| Partially vaccinated | Immunocompromised | 3 | mRNA | mRNA | mRNA | -- | -- | -- | 0-6 days | Excluded |
| Partially vaccinated | Immunocompromised | 1 | J&J[c] | -- | -- | -- | -- | -- | N/A | Excluded |
| Partially vaccinated | Immunocompromised | 2 | J&J | Any | -- | -- | -- | -- | 0-6 days | Excluded |
| Completed primary series | Immunocompetent | 1 | J&J | -- | -- | -- | -- | -- | ≥14 days | 98/3,181 (3%) |
| Completed primary series | Immunocompetent | 2 | J&J | Any | -- | -- | -- | -- | 0-6 days | 12/3,181 (0%) |
| Completed primary series | Immunocompetent | 2 | mRNA | mRNA | -- | -- | -- | -- | ≥14 days | 711/3,181 (22%) |
| Completed primary series | Immunocompetent | 3 | mRNA | mRNA | Any | -- | -- | -- | 0-6 days | 40/3,181 (1%) |
| Completed primary series | Immunocompromised | 2 | J&J | Any | -- | -- | -- | -- | ≥7 days | 0/3,181 (0%) |
| Completed primary series | Immunocompromised | 3 | J&J | Any | Any | -- | -- | -- | 0-6 days | 0/3,181 (0%) |
| Completed primary series | Immunocompromised | 3 | mRNA | mRNA | mRNA | -- | -- | -- | ≥7 days | 465/3,181 (15%) |
| Completed primary series | Immunocompromised | 4 | mRNA | mRNA | mRNA | Any | -- | -- | 0-6 days | 0/3,181 (0%) |
| Primary series + booster dose | Immunocompetent | 2 | J&J | Any | -- | -- | -- | -- | ≥7 days | 80/3,181 (3%) |
| Primary series + booster dose | Immunocompetent | 3 | J&J | Any | Any | -- | -- | -- | 0-6 days | 0/3,181 (0%) |
| Primary series + booster dose | Immunocompetent | 3 | mRNA | mRNA | Any | -- | -- | -- | ≥7 days | 710/3,181 (22%) |
| Primary series + booster dose | Immunocompetent | 4 | mRNA | mRNA | Any | Any | -- | -- | 0-6 days | 3/3,181 (0%) |
| Primary series + booster dose | Immunocompromised | 3 | J&J | Any | Any | -- | -- | -- | ≥7 days | 1/3,181 (0%) |
| Primary series + booster dose | Immunocompromised | 4 | J&J | Any | Any | Any | -- | -- | 0-6 days | 0/3,181 (0%) |
| Primary series + booster dose | Immunocompromised | 4 | mRNA | mRNA | mRNA | Any | -- | -- | ≥7 days | 4/3,181 (0%) |
| Primary series + booster dose | Immunocompromised | 5 | mRNA | mRNA | mRNA | Any | Any | -- | 0-6 days | 0/3,181 (0%) |

[a] Immunocompromising conditions included: active solid organ cancer (active cancer defined as treatment for the cancer or newly diagnosed cancer in the past 6 months), active hematologic cancer (such as leukemia, lymphoma, or myeloma), HIV infection without AIDS, AIDS, congenital immunodeficiency syndrome, previous splenectomy, previous solid organ transplant, immunosuppressive medication, systemic lupus erythematosus, rheumatoid arthritis, psoriasis, scleroderma, or inflammatory bowel disease, including Crohn's disease or ulcerative colitis.

[b] mRNA vaccine refers to either BNT162b2 (Pfizer-BioNTech) or mRNA-1273 (Moderna)

[c] J&J refers to Ad26.COV2 (Johnson & Johnson [Janssen])

Table S4. Booster dose eligibility.

Timeline of eligibility for additional and booster doses of COVID-19 vaccines

| **Date** | **Authorization** |
|---|---|
| August 13, 2021 | Immunocompromised patients recommended to receive an additional primary series dose of Pfizer-BioNTech or Moderna vaccine, following initial series of 2 doses of mRNA<br><br>[Centers for Disease Control and Prevention. ACIP Presentation Slides: August 13, 2021 Meeting. Updated August 13, 2021. Accessed April 15, 2022 https://www.cdc.gov/vaccines/acip/meetings/slides-2021-08-13.html)] |
| September 22, 2021 | Individuals ≥65 years, those at high risk of COVID-19-associated hospitalization, and immunocompromised patients recommended to receive a booster dose of Pfizer-BioNTech, following primary series of mRNA or Janssen (Johnson & Johnson)<br><br>[Centers for Disease Control and Prevention. ACIP Presentation Slides: Sept 22-23, 2021 Meeting. Updated September 23, 2021. Accessed April 15, 2022, https://www.cdc.gov/vaccines/acip/meetings/slides-2021-09-22-23.html] |
| October 21, 2021 | Moderna and Janssen (Johnson & Johnson) recommended for use as booster dose products for immunocompromised, ≥65 years, and high-risk individuals<br><br>[Mbaeyi S, Oliver SE, Collins JP, et al. The Advisory Committee on Immunization Practices' Interim Recommendations for Additional Primary and Booster Doses of COVID-19 Vaccines - United States, 2021. *MMWR Morb Mortal Wkly Rep*. Nov 05 2021;70(44):1545-1552. doi:10.15585/mmwr.mm7044e2] |
| November 19, 2021 | All adults (≥18 years) recommended to receive a booster dose of Pfizer-BioNTech or Moderna vaccine, following primary series of 2 doses of mRNA or 1 dose of Janssen (Johnson & Johnson)<br><br>[Centers for Disease Control and Prevention. ACIP Presentation Slides: November 19, 2021 Meeting. Updated November 19, 2021. Accessed April 15, 2022, https://www.cdc.gov/vaccines/acip/meetings/slides-2021-11-19.html] |
| March 29, 2022 | Individuals ≥50 years or immunocompromised allowed to receive a second booster dose of Pfizer-BioNTech or Moderna vaccine, following primary series and first booster dose<br><br>[Centers for Disease Control and Prevention. CDC Recommends Additional Boosters for Certain Individuals. Updated March 29, 2022. Accessed April 15, 2022, https://www.cdc.gov/media/releases/2022/s0328-covid-19-boosters.html] |
| May 19, 2022 | Individuals ≥50 years or immunocompromised recommended to receive a second booster dose of Pfizer-BioNTech or Moderna vaccine, following primary series and first booster dose<br><br>[Centers for Disease Control and Prevention. CDC Strengthens Recommendations and Expands Eligibility for COVID-19 Booster Shots. Updated May 19, 2022. Accessed May 20, 2022, https://www.cdc.gov/media/releases/2022/s0519-covid-booster-acip.html] |

Table S5. Healthy volunteers in antibody evaluation.

Characteristics of healthy volunteers in the analysis of anti-SARS-CoV-2 antibody concentrations after COVID-19 booster doses.

| **Characteristic** | **BNT162b2 three dose series (n=33)** | **mRNA-1273 three dose series (n=16)** | **Ad26.COV2 two dose series (n=8)** | **Ad26.COV2 one dose, then mRNA booster (n=6)** |
|---|---|---|---|---|
| Age, median (IQR), years | 48 (32, 57.75) | 43 (26.5, 67.5) | 55 (51, 56.75) | 57.5 (36.75, 63.25) |
| Female sex, no. (%) | 21(63.6) | 8 (50) | 3 (37.5) | 4 (66.7) |
| Male sex, no. (%) | 12 (36.4) | 8 (50) | 5 (62.5) | 2 (33.3) |
| Race/ethnicity, no. (%) | | | | |
| Non-Hispanic White | 25 (75.8) | 14 (87.5) | 8 (100) | 5 (83.3) |
| Non-Hispanic Black | 6 (18.2) | 2 (12.5) | 0 (0) | 0 (0) |
| Hispanic | 2 (6.1) | 0 (0) | 0 (0) | 0 (0) |
| Non-Hispanic Asian | 0 (0) | 0 (0) | 0 (0) | 0 (0) |
| Non-Hispanic American Indian or Alaska Native | 0 (0) | 0 (0) | 0 (0) | 1 (16.7) |
| Non-Hispanic Native Hawaiian or Other Pacific Islander | 0 (0) | 0 (0) | 0 (0) | 0 (0) |
| Non-Hispanic, all other races | 0 (0) | 0 (0) | 0 (0) | 0 (0) |
| Time between full vaccination and receipt of booster, median (IQR), days | 240 (219.5, 284) | 233 (208, 255.5) | 233 (219, 244.75) | 230.5 (219.25, 238) |
| Time between receipt of booster and post-booster blood draw, median (IQR), days | 23 (18, 29.5) | 19 (16.5, 30) | 25 (22.5, 29.5) | 25.5 (18, 32.25) |

Table S6. Anti-SARS-CoV-2 antibody concentrations.

Anti-SARS-CoV-2 antibody concentrations among healthy volunteers after full COVID-19 vaccination, before a first booster dose, and after a first booster dose. Results for individual patients are shown in Figure 4.

(A) All participants with antibody measurements before and after booster dose.

| **COVID-19 vaccine series** | **n** | **Anti-RBD concentration (BAU/ml), geometric mean (95% CI)** | | **Anti-spike concentration (BAU/ml), geometric mean (95% CI)** | |
|---|---|---|---|---|---|
| | | Before booster dose | 2-6 weeks after booster dose | Before booster dose | 2-6 weeks after booster dose |
| BNT162b2 three dose series | 33 | 199.3 (142.3,279.3) | 8708.7 (6547.6, 11583.2) | 156.7 (116.5, 210.8) | 5897.9 (4446.4, 7823.3) |
| mRNA-1273 three dose series | 16 | 467.0 (293.8, 742.3) | 10479.0 (7765.2, 14141.4) | 340.3 (216.1, 536.1) | 6986.2 (5058.2, 9649.2) |
| Ad26.COV2 two dose series | 8 | 51.1 (27.1, 96.5) | 961.2 (431.7, 2140.1) | 37.1 (20.9, 66.1) | 702.2 (318.2, 1549.8) |
| Ad26.COV2 one dose, then mRNA booster | 6 | 38.6 (7.0, 213.1) | 3781.1 (1517.3, 9422.3) | 29.9 (5.82, 154.2) | 2669.9 (1093.3, 6519.9) |

Abbreviations: BAU = binding antibody units

(B) Subset of participants with antibody measurements before and after a booster dose who also had antibody measurements after full vaccination.

| **COVID-19 vaccine series** | **n** | **Anti-RBD concentration (BAU/ml), geometric mean (95% CI)** | | | **Anti-spike concentration (BAU/ml), geometric mean (95% CI)** | | |
|---|---|---|---|---|---|---|---|
| | | 2-6 weeks after full vaccination | Before booster dose | 2-6 weeks after booster dose | 2-6 weeks after full vaccination | Before booster dose | 2-6 weeks after booster dose |
| BNT162b2 three dose series | **9** | 3078.7 (1554.6, 6097.0) | 246.9 (116.6, 523.2) | 10484.9 (6380.4, 17229.8) | 2989.7 (1568.8, 5697.6 | 205.9 (126.0, 336.3) | 7260.2 (3842.43, 13717.9) |
| mRNA-1273 three dose series | **7** | 4912.7 (3283.3, 7350.8) | 895.9 (558.3, 1437.6) | 12856.2 (7205.7, 22937.9) | 3487.8 (2199.7, 5530.2) | 573.7 (343.5, 958.3) | 8257.1 (4558.2, 14957.6) |
| Ad26.COV2 two dose series | **3** | 12.76 (4.7, 34.5) | 26.3 (12.5, 55.7) | 501.2 (35.3, 7110.4) | 11.79 (3.31, 41.9) | 22.2 (4.9, 100.8) | 360.4 (27.9, 4655.3) |
| Ad26.COV2 one dose, then mRNA booster | **2** | 77.6 (66.2, 91.0) | 20.62 (0, 453557.5) | 2361.2 (0.1, 1049821470 | 116.6 (3.09, 4389.5) | 18.6 (0, 583040.7) | 1646.8 (0.1, 46373002) |

Abbreviations: BAU = binding antibody units

Appendix C: Supplementary Figures

Figure S1. Participant flow diagram.

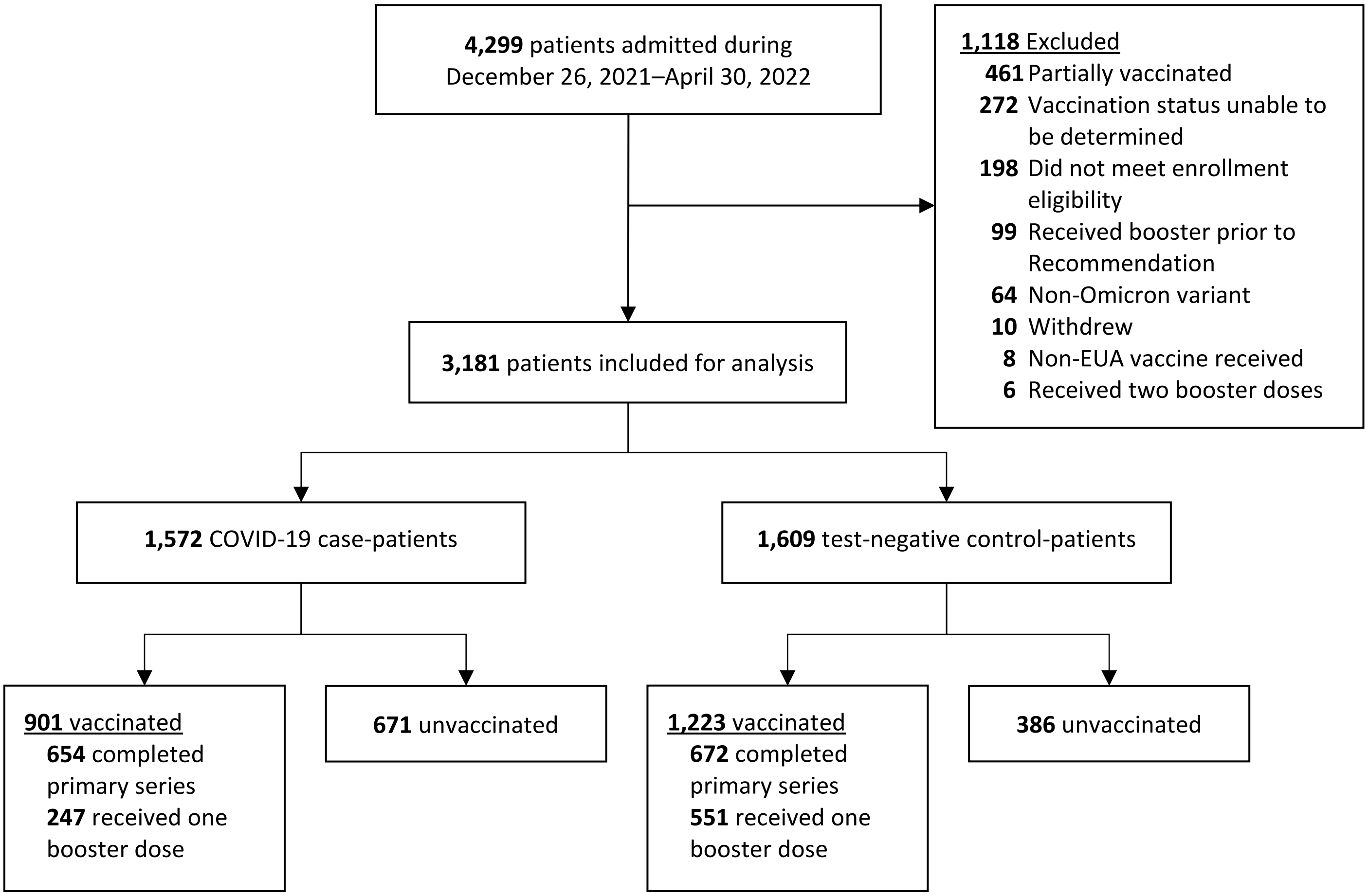
